# Natural Language Processing to Identify Patients with Cognitive Impairment

**DOI:** 10.1101/2022.02.16.22271085

**Authors:** Khalil I Hussein, Lili Chan, Tielman Van Vleck, Kelly Beers, Monica R Mindt, Michael Wolf, Laura M. Curtis, Parul Agarwal, Juan Wisnivesky, Girish N. Nadkarni, Alex Federman

## Abstract

**INTRODUCTION:** Early detection of patients with cognitive impairment may facilitate care for individuals in this population. Natural language processing (NLP) is a potential approach to identifying patients with cognitive impairment from electronic health records (EHR).

**METHODS:** We used three machine learning algorithms (logistic regression, multilayer perceptron, and random forest) using clinical terms extracted by NLP to predict cognitive impairment in a cohort of 199 patients. Cognitive impairment was defined as a mini-mental status exams (MMSE) score <24.

**RESULTS:** NLP identified 69 (35%) patients with cognitive impairment and ICD codes identified 44 (22%). Using MMSE as a reference standard, NLP sensitivity was 35%, specificity 66%, precision 41%, and NPV 61%. The random forest method had the best test parameters; sensitivity 95%, specificity 100%, precision 100%, and NPV 97%

**DISCUSSION:** NLP can identify adults with cognitive impairment with moderate test performance that is enhanced with machine learning.

## Introduction

Cognitive impairment is common among Americans over the age of 65, with an estimated prevalence as high as 9% for dementia and 28% for mild cognitive impairment in some populations.^1, 2^ Further, the absolute number of individuals with cognitive difficulties will continue to rise as the U.S. population ages.^3^ Early detection of patients with cognitive impairment may enable primary care providers to facilitate care and care management and improve outcomes for individuals in this population.^4^ Yet systematically identifying patients with cognitive impairment in clinical settings has proven to be logistically difficult.^5^ For example, validated tools for cognitive impairment screening, like the mini Mental State Exam (MMSE) and the Montreal Cognitive Assessment (MoCA), are infrequently used in clinical care, possibly owing to the competing demands of management of multimorbidity in the primary care of older adults.^6-11^ Therefore, a method to efficiently and accurately identify mild cognitive impairment that minimizes primary care provider involvement is necessary.

The rise of healthcare information technology and big data analytics present potential new opportunities to circumvent the existing challenges of identifying patients with cognitive impairment. Electronic health records (EHR) hold an enormous quantum of data beyond traditionally used discrete data elements, like International Classification of Diseases (ICD) diagnosis codes. Free text documentation by clinicians and other members of the health system may hold information about cognitive abilities for the individual, ranging from the nuanced (e.g., “the patient forgets”) to the overt (e.g., “family is concerned about dementia”). Such data can be leveraged using advanced informatics approaches for cognitive impairment research.^12^ Natural language processing (NLP) is such an approach. NLP can be used to process large volumes of free text in clinical documentation and convert it into discrete data elements suitable for quantitative analysis.^13^ Machine learning can then be applied to the data elements extracted through NLP to create precise prediction models when a standard measure of cognition is available. In this study, we used NLP and machine learning to identify patients with cognitive impairment using EHR data from an academic medical center and a neurocognitive assessment as the reference standard.

## Methods

### Study Population

Data for this study were obtained from multiple sources. While all patients were part of the BioMe Biobank, patient interview data was obtained from two cohort studies.^14^ The data from these sources were used to characterize study patients and provide a reference standard for cognitive impairment. The cohort studies were conducted in primary care and pulmonary practices in New York City and Chicago, included older adults (ages ≥50 years) with chronic obstructive pulmonary disease (COPD) or asthma, and excluded individuals with dementia based on ICD coding in the electronic record. Patients with diagnosed dementia were excluded since the primary goal of this study was to identify patients with previously undiagnosed cognitive impairment. In both studies, baseline assessments of cognition were conducted by research assistants who were formally trained and supervised in the administration of neuropsychological assessment, including the MMSE, by a research psychologist.^15, 16^

The Bio*Me* Biobank is a prospective registry of patients recruited from primary care and subspecialty clinics in the Mount Sinai Health System. Participants consent to the use of their EHR data for biomedical research and the Mount Sinai Institutional Review Board approved the BioME protocols. For the purpose of the present study, we included patients in the cohort studies who had also consented to BioMe participation, and linked their EHR data with cohort study survey data. We retrieved all clinical notes of participants available from the EHR up to December 31, 2017.

### Study Design

#### Reference Standards for Cognitive Impairment

The primary reference standard was the mini-Mental State Exam, which was administered to subjects in both cohort studies at baseline, 12 months, and 24 months. Research assistants were formally trained and supervised in the administration of neuropsychological assessment by a research psychologist from the Mount Sinai School of Medicine Alzheimer’s Disease Research Center. For Spanish-speaking patients, the MMSE was administered in Spanish. We defined cognitive impairment as an unadjusted MMSE score of <24 and used data from the most recent assessment.^17^

In a set of secondary analyses, we used physician review of the chart to determine whether there was documented evidence of cognitive impairment. We randomly selected 25 patients who were positive for cognitive impairment by NLP and 25 who were negative. Chart review was independently performed by 2 physicians who were blinded to the results of the NLP analyses and to the ICD codes. Disagreements between the 2 physicians were adjudicated by a third physician. Each reviewer read through all available notes in the EHR. Patients were considered to have cognitive impairment if reviewers found documentation of patient or patient family member complaints of forgetfulness, difficulty learning new things, concentrating, or making decisions to the point of interfering with their everyday life. This could be intermittent or related to a transient condition, such as delirium. Additionally, if a patient was referred to a specialist for dementia workup, reviewers considered this to be positive for cognitive impairment. However, patients forgetting to take medications or to bring in blood pressure/blood sugar logs were not considered to be cognitively impaired, nor were patients who developed altered mental status or cognitive decline while under inpatient hospice care.

#### Natural Language Processing

NLP was used to parse all available progress notes and discharge summaries. When using MMSE as a reference only notes from a 24-month period, between 12 months prior to and 12 months after the most recent MMSE administration was used. Since the MMSE was obtained at a defined time, this time restriction allows for inclusion of only notes that reflect the provider assessment of the patients’ cognitive function around the time of the MMSE. This avoids including notes from patients who may have developed cognitive impairment after the MMSE. However, when we compared the NLP algorithm to manual chart review and ICD codes, since the entire chart was reviewed, there were no time restrictions to the notes for NLP querying. We excluded radiology reports and pathology reports as text from these note types are generally devoid of assessments of cognition. The NLP program matched words and phrases in EHR free-text documentation to clinical terms of the Systematized Nomenclature of Medicine – Clinical Terms (SNOMED CT). SNOMED is a comprehensive healthcare terminology consisting of hierarchies of concepts, with parent terms encompassing specific concepts (child terms).^18^ Two physicians independently reviewed available SNOMED CT for terms associated with cognitive impairment to be used for NLP querying. The SNOMED CT parent and child terms included in the query are diagrammed in eFigure 1.

For each of these terms a query was created to identify instances of the concept documented as present in the clinical record and determined whether it represented the subject of record (the patient vs. a family member) and its temporality (current or past). We defined cognitive impairment by NLP as identification of ≥1 term in the medical record linked to cognitive impairment by SNOMED CT. Cognitive impairment was not considered present when negation terms, e.g., not or no, were used in the same sentence or when cognitive impairment was mentioned in the context of family history. NLP was performed using CLiX NLP (Clinithink, London, UK).

After first use of NLP to identify patients with cognitive impairment, we conducted a manual review of 50 randomly selected charts to identify and correct inaccuracies in the NLP strategy that could lead to false positive and negative results. For example, NLP labeled one patient as having cognitive impairment when it recognized the “mCi” abbreviation used for “millicurie” (a unit of radioactivity) as indicative of mild cognitive impairment.

#### Identifying Cognitively Impaired Patients by ICD Codes

We also used ICD 9 and 10 codes to identify patients with cognitive impairment (eTable 1).^19, 20^ The codes we used included 290.x (dementias, including senile and vascular), 294.x (persistent mental disorders due to conditions classified elsewhere, e.g., amnestic disorder in conditions classified elsewhere, dementia unspecified), F01.x (vascular dementia), and F01.x (unspecified dementia).

**Table 1:** Demographics

| Characteristic |  | Total<br>(n=199)<br>n (%) | MMSE<24<br>(n=79)<br>n (%) | MMSE≥24<br>(n=120)<br>n (%) | p-value |
| --- | --- | --- | --- | --- | --- |
| Age |  |  |  |  |  |
|  | 55-64 | 88 (44.2%) | 25 (31.7%) | 63 (52.5%) | 0.008 |
|  | 65-70 | 58 (29.2%) | 31 (39.2%) | 27 (22.5%) |  |
|  | ≥71 | 53 (26.6%) | 23 (29.1%) | 30 (25.0%) |  |
| Female |  | 149 (74.9%) | 56 (70.9%) | 93 (77.5%) | 0.29 |
| Married/Living with Partner |  | 44 (22.1%) | 17 (21.5%) | 27 (22.5%) | 0.87 |
| Race |  |  |  |  |  |
|  | White | 29 (14.6%) | 7 (8.9%) | 22 (18.3%) | 0.27 |
|  | Black | 56 (28.1%) | 22 (27.8%) | 34 (28.3%) |  |
|  | Hispanic | 106 (53.3%) | 47 (59.5%) | 59 (49.2%) |  |
|  | Other | 8 (4.0%) | 3 (3.8%) | 5 (4.2%) |  |
| Monthly Income |  |  |  |  |  |
| | \$0-\$750 | 100 (50.8%) | 38 (48.1%) | 62 (51.7%) | 0.63 |
| | \$751-\$1350 | 62 (31.5%) | 28 (35.4%) | 34 (28.3%) | |
| | \$1351-\$3000 | 23 (11.6%) | 8 (10.1%) | 15 (12.5%) | |
| | >\$3000 | 6 (3.1%) | 1 (1.3%) | 5 (4.2%) | |
|  | Refused/Don't Know | 8 (4.0%) | 4 (5.1%) | 4 (3.3%) |  |
| Education |  |  |  |  |  |
|  | Less than 12 years | 91 (45.7%) | 50 (63.3%) | 41 (34.2%) | <0.001 |
|  | High school graduate | 42 (21.1%) | 12 (15.2%) | 30 (25.0%) |  |
|  | Some college | 35 (17.6%) | 9 (11.4%) | 26 (21.7%) |  |
|  | College degree or higher | 30 (15.1%) | 7 (8.9%) | 23 (19.2%) |  |
|  | Refused/Don't Know | 1 (0.5%) | 1 (1.3%) | 0 (0.0%) |  |
| General Health Rating |  |  |  |  |  |
|  | Excellent/Very Good | 29 (14.6%) | 6 (7.6%) | 23 (19.2%) | 0.003 |
|  | Good | 53 (26.6%) | 15 (19.0%) | 38 (31.7%) |  |
|  | Fair/Poor | 117 (58.8%) | 58 (73.4%) | 59 (49.2%) |  |
| Assistance with ADLs |  |  |  |  |  |
|  | No help | 123 (61.8%) | 42 (53.1%) | 81 (67.5%) | 0.08 |
|  | Help needed | 74 (37.2%) | 36 (45.6%) | 38 (31.7%) |  |
|  | Refused/Don't Know | 2 (1.0%) | 1 (1.3%) | 1 (0.8%) |  |
| Origin Country/Territory |  |  |  |  |  |
|  | United States | 103 (51.8%) | 33 (41.8%) | 70 (58.3%) | 0.02 |
|  | Puerto Rico | 73 (36.7%) | 34 (43.0%) | 39 (32.5%) |  |
|  | Dominican Republic | 9 (4.5%) | 7 (8.9%) | 2 (1.7%) |  |
|  | Other | 14 (7.0%) | 5 (6.3%) | 9 (7.5%) |  |
| Preferred Language |  |  |  |  |  |
|  | English | 147 (73.9%) | 46 (58.3%) | 101 (84.2%) | <0.001 |
|  | Spanish | 52 (26.1%) | 33 (41.7%) | 19 (15.8%) |  |

#### Machine Learning Strategy

In order to develop a machine learning approach to predicting cognitive impairment from free text data, all SNOMED terms were extracted from every available progress note for all patients for the 24 month period of observation. Each data element matched to a SNOMED clinical term has 4 features (temporality, association, subject relationship, finding or presence of the condition). An example is presented in eFigure 2 for the term “forgetful”. While an asset when trying to describe patient features in great detail, the granularity of textual data parsed by NLP complicates the correlation of patients with similar traits when sample sizes (number of patients) are small. To resolve this complexity, we eliminated parts of the SNOMED expression that were unnecessary for analysis such as redundant modifiers. As the SNOMED expressions alone do not capture the logical hierarchy of SNOMED, we also walked up the SNOMED hierarchy and created additional features for relevant children concepts. Thus, for the parent term “cognitive impairment,” the additional features were created to represent the children concepts of forgetful, memory impairment, memory finding, cognitive function finding, mental state, behavior and/or psychosocial function finding, and impaired cognition. We then trained and tested machine learning classifiers using only the condensed output from our NLP algorithm to predict whether the patient would have MMSE score <24. Classification methods included logistic regression, multilayer perceptron (MLP) a feedforward neural network^21^, and random forest.^22^ We performed 100-fold cross validation. Models were trained on a varying number of top features, according to K-Means correlation with MMSE <24. All machine learning procedures were performed in Python, using the standard scikit-learn package.

#### Statistical Analysis

We assessed performance of NLP, ICD, and the combination of NLP and/or ICD codes for identification of patients with cognitive impairment by calculating the sensitivity, specificity, precision (also knowns as positive predictive value), negative predictive value (NPV), and F1 scores, using MMSE scores and determination by manual chart review as the primary and secondary reference standards, respectively. We performed comparison of categorical demographic variables by cognitive impairment status using Chi Square and Fisher Exact tests and t-tests for continuous variables. We calculated kappa statistic for inter-rater agreement for manual chart review using SAS Macro MAGREE. All statistics were calculated using SAS version 9.4 (SAS Institute, Cary, NC).

## Results

### Subject Characteristics

We linked EHR and cognitive assessment data for 199 patients. The average age of patients was 68±7. They were predominantly female (75%), Hispanic (53%), and low income (<$750 per month, 51%); 46% had completed less than 12 years of formal education (Table 1).

MMSE score was less than 24 for 79 (40%) patients and was more common with increasing age, lower education, poorer general health, and Spanish language preference.

### Performance of NLP and ICD Codes with MMSE Assessment as the Reference Standard

NLP identified 69 (35%) patients as having cognitive impairment and ICD codes identified 44 (22%). Sensitivity of NLP for detection of cognitive impairment was low, 0.35 (95% CI 0.25-0.47), while specificity was moderate, 0.66 (95% CI 0.57-0.74) (Table 2). Use of ICD 9 and 10 codes to detect cognitive impairment performed similarly (sensitivity, 0.24 (95% CI 0.13-0.59); specificity, 0.79 (95% CI 0.6-0.91). Negative and positive predictive and F1 values were also similar for the two strategies (Table 2). Combining NLP and ICD codes into a single diagnostic strategy did not meaningfully alter test performance compared to either approach alone: sensitivity, 0.41 (95% CI 0.3-0.52); specificity, 0.63 (95% CI 0.54-0.72).

**Table 2:** NLP and ICD9/10 compared with MMSE (inclusive of notes +/-1 year of MMSE)

|  | NLP | ICD | NLP/ICD |
| --- | --- | --- | --- |
| Sensitivity | 0.35 (0.25-0.47) | 0.24 (0.13-0.59) | 0.41 (0.30-0.52) |
| Specificity | 0.66 (0.57-0.74) | 0.79 (0.60-0.91) | 0.63 (0.54-0.72) |
| Precision | 0.41 (0.29-0.53) | 0.43 (0.19-0.75) | 0.42 (0.31-0.54) |
| NPV | 0.61 (0.52-0.69) | 0.61 (0.50-0.82) | 0.62 (0.53-0.70) |
| F1 Score | 0.38 | 0.31 | 0.41 |

### Performance of NLP and ICD Codes with Manual Chart Review as the Reference Standard

Agreement between reviewers for chart review was high (kappa 0.78, 95% CI 0.62-0.94). Sensitivity of NLP for cognitive impairment as determined by manual chart review was high, 0.96 (95% CI 0.75-1), specificity was moderate, 0.68 (95% CI 0.5-0.82), and precision was moderate 0.52 (95% CI 0.31-0.72) (Table 3). ICD 9 and 10 codes for cognitive impairment by manual chart review had moderate sensitivity, 0.77 (95% CI 0.46-0.95), high specificity, 0.92 (95% CI 0.78-0.98), and moderate precision 0.77 (95% CI 0.46-0.95). NPV and F1 scores were similar for the two strategies (Table 3). The combination of NLP and ICD codes as a single diagnostic strategy did not substantially change test performance: sensitivity 0.96 (95% CI 0.75-1), specificity 0.7 (95% CI 0.5-0.82), precision 0.52 (95% CI 0.31-0.72), NPV 0.98 (95% CI 0.86-1), and F1 score of 0.68.

**Table 3:** NLP and ICD9/10 compared with Manual Chart Review (inclusive of all notes)

|  | NLP | ICD | NLP/ICD |
| --- | --- | --- | --- |
| Sensitivity | 0.96 (0.75-1.00) | 0.77 (0.46-0.95) | 0.96 (0.75-1.00) |
| Specificity | 0.68 (0.50-0.82) | 0.92 (0.78-0.98) | 0.68 (0.50-0.82) |
| Precision | 0.52 (0.31-0.72) | 0.77 (0.46-0.95) | 0.52 (0.31-0.72) |
| NPV | 0.98 (0.86-1.00) | 0.92 (0.78-0.98) | 0.98 (0.86-1.00) |
| F1 Score | 0.68 | 0.77 | 0.68 |

### Machine Learning with NLP terms for Identification of Cognitive Impairment

Application of machine learning to NLP-identified terms resulted in substantial improvements in identification of patients with cognitive impairment with respect to the MMSE reference standard (Table 4). Of the three classifiers tested, the Random Forest method performed best, though only slightly better than the MLP neural net. The Random Forest approach yielded a sensitivity of 0.95, specificity 1.00, precision 1.00, NPV 0.97, F1 score of 0.98 with overall AUC 0.98. For Supervised Neural Network (MLP Classifier) approach, sensitivity was 0.94, specificity 1.00, precision 1.00, NPV 0.96, F1 score of 0.97 with overall AUC 0.97. Lastly, for the logistic regression approach sensitivity was 0.63, specificity 0.98, precision 0.95, NPV 0.79, F1 score of 0.76 with overall AUC 0.80.

**Table 4:** Machine Learning Applied to NLP extracted terms compared with MMSE

|  | Logistic Regression | MLP | Random Forest |
| --- | --- | --- | --- |
| Sensitivity | 0.63 | 0.94 | 0.95 |
| Specificity | 0.98 | 1.00 | 1.00 |
| Precision | 0.95 | 1.00 | 1.00 |
| NPV | 0.79 | 0.96 | 0.97 |
| F1 Score | 0.76 | 0.97 | 0.98 |
| AUC | 0.80 | 0.97 | 0.98 |

## Discussion

In this study, we used NLP to identify patients with cognitive impairment from EHR documentation and found that it had modest test performance in relation to the MMSE, a standardized assessment. NLP performed similarly to ICD codes when using MMSE as the reference standard but had better performance when using manual review as the reference standard. However, applying machine learning approaches to the concepts extracted by NLP greatly improved test performance.

Our study builds on prior literature aimed at detection of patients with cognitive impairment. Multiple studies have utilized NLP to analyze patient speech patterns to identify cognitive impairment.^23-25^ However, this approach requires prospective collection of recordings or transcription of patient-physician visits and may be logistically challenging in clinical practice. Only one study has used NLP with EHR data.^20^ Reuben et al used medications, ICD codes, and NLP to identify patients with dementia and compared it to physician manual chart review. Their NLP algorithm only included terms for “dementia” or “neurodegenerative” without negation terms or family history markers. We used a more complex NLP algorithm that was based on SNOMED terms and hierarchy, therefore including a more extensive list of terms for querying. This may have contributed to the higher sensitivity we found compared to that reported by Reuben et al. Additionally, our main comparison was between NLP and an objective cognitive assessment with the MMSE, rather than chart review.

As the proportion of older adults in the US population increases, the number of patients with cognitive impairment is also increasing. Cognitive impairment puts a strain on the US healthcare system as it is a major risk factor for hospital admission and readmissions, and a major contributor of healthcare costs among older adults.^26-29^ Additionally, cognitive impairment negatively impacts an individual’s life by affecting their ability to self-manage chronic diseases, as a risk factor for functional status decline, and contributes to the development of depression and other chronic health problems.^30-33^ Despite this, cognitive impairment is under-recognized with only 8-28% of older adults every being screened and on average 10 years between the appearance of early declines in cognitive function and a clinician diagnosis.

While our NLP and machine learning approach requires additional validation, since it is based on existing EHR, it can be easily implemented into EHR systems. One proposed method would be a clinical decision support system, which can notify patient’s providers when the algorithm identifies a patient with cognitive impairment and suggest potential actions to take such as referrals to neuropsychological testing or assessment for the need of a home health aide. This EHR-based intervention could most benefit patients who regularly interact with healthcare providers that typically do not screen existing clinical data in the chart for evidence of cognitive impairment. For example, an elderly patient who is frequently admitted to a hospital and regularly follows up with multiple specialists for non-cognitive concerns but rarely sees a primary care physician, geriatrician, psychiatrist, or neurologist could easily avoid formal cognitive screening despite risk factors. Because our proposed cognitive impairment detection tools would only use data already contained within the EHR, would only be visible to the patient’s providers, and would only suggest the initiation of the cognitive impairment diagnostic workup rather than attempt to establish the diagnosis itself, this intervention would not raise privacy or other ethical concerns.

Early detection of cognitive impairment is advocated by the US Department of Health and Human Services and the Alzheimer’s Foundation.^34, 35^ Early identification can allow for enrollment into programs to exclude reversible causes of dementia, cognitive intervention programs^36^, pharmacotherapy when appropriate, and identifying care coordination needs.^37^ Additionally, identification will enable care givers to receive support to alleviate the stress and burden associated with caring for people with cognitive impairment. Lastly, effective and efficient methods of identifying patients with cognitive impairment will allow for inclusion of these patients into clinical trials and cohort studies. Our NLP and machine learning approach is a scalable method that can facilitate identification given its high sensitivity, understanding that there is the potential for false positive given the lower specificity. As with any screening test, there can be false negatives and false positives. A false negative will lead to a delay in a patient with cognitive impairment receiving appropriate referrals for care, while a false positive will lead to unnecessary additional testing and can cause patient’s unneeded stress. Therefore, validation of our results are necessary before implementation.

In this study, patients who had lower MMSE scores were older, had less education, and preferred to communicate in Spanish. It has been well established that age, education, and language are associated with lower MMSE scores.^38^ Some researchers have performed age and education adjustment, which increases the sensitivity but decreases the specificity of the MMSE.^39^ Lastly, others have argued against using adjustment for education as there may be a true association between education level and cognitive impairment.^40^ Due to this uncertainty and that our study is a proof of concept that NLP can be used to identify patients with cognitive impairment, we chose to use the unadjusted cutoff of 24 consistent with mild impairment that is a widely accepted threshold for cognitive impairment in our analyses.

We chose to not include medications in the NLP or machine learning algorithm. Prior studies did not find that the addition of medications improved performance of their NLP or ICD algorithms for dementia identification.^20^ Authors cite the use of dementia medications for alternative diagnoses as a possible cause. We found a higher prevalence of cognitive impairment than the 15-25% that is reported in literature, which may be due to the inclusion of only older adults with asthma or COPD, two conditions having established associations with cognitive impairment in older adults.^41^ Differences between sensitivity of NLP when using MMSE as a standard and chart review as a standard may be influenced by the under diagnosis of cognitive impairment by providers and therefore not documented in the chart.^42^

This study has several limitations. First, the sample size was relatively small, which could have led to overfitting of the machine learning algorithm. We performed a 100-fold cross validation to reduce the risk of overfitting, but validation with data from a different health system is needed before conclusions can be drawn about the NLP and machine learning strategies we employed. As NLP can only identify cognitive impairment from what is in the EHR, it is dependent on the number of encounters and notes available. While there are many methods of identifying cognitive impairment, we only used the MMSE, which can be biased by a person’s education, primary language, and culture. Despite these limitations, this is the first study to use NLP and machine learning to identify patients with cognitive impairment using data from EHR.

## Conclusion

NLP can be used to identify adults with cognitive impairment with moderate test performance and greatly enhanced with the addition of machine learning. NLP and machine learning out performed ICD codes for identification of cognitive impairment. While additional validation in external datasets is necessary, this method provides for a scalable and high throughput method for identifying patients with cognitive impairment for more appropriate diagnostic testing, early treatment, and enrollment into research studies.

## Data Availability

All data produced in the present study are available upon reasonable request to the authors.

## Acknowledgements

Data, analytic methods, and study materials can be made available after review and approval by institutional regulatory bodies. The study was not preregistered in any public registry.

## Author Contributions

KIH, LC, TVV, KB, PA, JW, GNN, and AF were responsible for the conception, design, and acquisition, design, and interpretation of the data. KIH, LC, GNN, AF, MRM, MW, LMC assisted in the drafting and critical revision of the manuscript. All authors approve of the final version of the manuscript.

