## Supplemental for "Natural Language Processing to Identify Patients with Cognitive Impairment"

**eTable 1: List of ICD codes used for identification of cognitive impairment.**

| | |
|---|---|
| **ICD-9 CM** | |
| 290.0, 290.1, 290.2, 290.3, 290.4, 290.8, 290.9 | Senile dementia, uncomplicated; presenile dementia; senile dementia with delusional or depressive features; senile dementia with delirium; vascular dementia; other specified senile psychotic conditions; unspecified senile psychotic condition. |
| 291.1, 291.2 | Alcohol-induced persisting amnestic disorder; alcohol-induced persisting dementia |
| 292.82 | Drug-induced persisting dementia |
| 294.0, 294.10, 294.11, 294.20 | Amnestic disorder in conditions classified elsewhere; dementia in conditions classified elsewhere without behavioral disturbance, dementia in conditions classified elsewhere with behavioral disturbance; Dementia, unspecified, without behavioral disturbance; |
| 331.0, 331.82, 331.11, 331.19 | Alzheimer's disease; Dementia with lewy bodies; Pick's disease; Other frontotemporal dementia |
| **ICD-10 CM** | |
| A81.00 | Creutzfeldt-Jakob disease unspecified |
| E71.0, E75.2x, E75.23, E75.29 | Maple-syrup-urine disease; Other sphingolipidosis; Other sphingolipidosis Krabbe disease; Other sphingolipidosis Other sphingolipidosis |
| F01.x, F02.x, F03.x, F04, F10.26, F10.27, F10.96, F10.97, , F13.26, F13.27, F13.96, F13.97, F18.27, F18.97 | Vascular dementia without behavioral disturbance; Vascular dementia with behavioral disturbance; Dementia in other diseases classified elsewhere without behavioral disturbance; Dementia in other diseases classified elsewhere with behavioral disturbance; Unspecified dementia without behavioral disturbance; Amnestic disorder due to known physiological condition; Alcohol dependence with alcohol-induced persisting dementia; Alcohol use, unspecified with alcohol-induced persisting amnestic disorder; Alcohol use, unspecified with alcohol-induced persisting dementia; Sedative, hypnotic or anxiolytic dependence with sedative, hypnotic or anxiolytic-induced persisting amnestic disorder; Sedative, hypnotic or anxiolytic dependence with sedative, hypnotic or anxiolytic-induced persisting dementia; Sedative, hypnotic or anxiolytic use, unspecified with sedative, hypnotic or anxiolytic-induced persisting amnestic disorder; Sedative, hypnotic or anxiolytic use, unspecified with sedative, hypnotic or anxiolytic-induced persisting dementia; Inhalant dependence with inhalant-induced dementia; Inhalant dependence with inhalant-induced persisting dementia |
| G10, G20, G30.x, G23.1, G31.X, G31.85 | Huntington's disease; Parkinson’s disease; Alzheimer's disease, unspecified; Progressive supranuclear ophthalmoplegia [Steele-Richardson- |

| | |
|---|---|
| | Olszewski]; Other degenerative diseases of nervous system, not elsewhere classified; Frontotemporal dementia; Pick's disease; Other frontotemporal dementia; Senile degeneration of brain, not elsewhere classified; Degeneration of nervous system due to alcohol; Other specified degenerative diseases of nervous system; Alpers disease; Dementia with Lewy bodies; Mild cognitive impairment, so stated; Corticobasal degeneration; Other specified degenerative diseases of nervous system; Degenerative disease of nervous system, unspecified |
| R41.x | Other symptoms and signs involving cognitive functions and awareness; Disorientation, unspecified; Anterograde amnesia; Retrograde amnesia; Other amnesia; Neurologic neglect syndrome; Other symptoms and signs involving cognitive functions and awareness; Age-related cognitive decline; Altered mental status, unspecified; Borderline intellectual functioning; Other specified cognitive deficit; Attention and concentration deficit; Cognitive communication deficit; Visuospatial deficit; Psychomotor deficit; Frontal lobe and executive function deficit; Other symptoms and signs involving cognitive functions and awareness; Unspecified symptoms and signs involving cognitive functions and awareness |

**eFigure 1: Clusters of SNOMED codes queried for including CI, dementia, movement disorders and degenerative brain disorders**

### Cognitive Impairment Query

#### Cognitive Impairment

- Age-related cognitive decline
- Cognitive changes due to organic disorder
- Cognitive deficit in attention
- Cognitive impairment co-occurrent and due to primary psychotic disorder
- Cognitive impairment due to toxicity of substance
- Dissociative neurological symptom disorder co-occurrent with cognitive symptoms
- Disturbance of cognitive learning
- Impaired environmental interpretation syndrome
- Impaired executive functioning
- Lack of thinking ability
- Memory impairment
- Minimal cognitive impairment
- Moderate cognitive impairment
- Severe cognitive impairment

#### Dementia

- Alzheimer's disease
- Delirium co-occurrent with dementia
- Dementia associated with another disease
- Dementia caused by heavy metal exposure
- Dementia caused by toxin
- Dementia co-occurrent with human immunodeficiency virus infection
- Dementia following injury caused by exposure to ionising radiation
- Dementia in remission
- Dementia of frontal lobe type
- Dementia with behavioural disturbance
- Dialysis dementia
- Drug-induced dementia (disorder)
- General paresis – neurosyphilis
- Mixed dementia
- Parkinson-dementia complex of Guam
- Parkinsonism with dementia of Guadeloupe
- Patchy dementia
- Predominantly cortical dementia
- Presenile dementia
- Primary degenerative dementia
- Rapidly progressive dementia
- Semantic dementia
- Senile and presenile organic psychotic conditions
- Senile dementia
- Subcortical dementia

#### Movement Disorder

- Diffuse Lewy Body Dementia
- Huntington disease-like 2
- Huntington disease-like syndrome
- Huntington's chorea
- Parkinson's disease
- Pallidoluysian degeneration
- Pallidonigral degeneration
- Pallidonigroluysian degeneration
- Pallidonigrospinal degeneration
- Pallidopontonigral degeneration

#### Degenerative Brain Disorder

- Cerebral degeneration
- Cystic degeneration of brain
  Degenerative brain disorder caused by alcohol
- Epilepsy co-occurrent and due to degenerative brain disorder
- GRN-related frontotemporal dementia
- Prion disease
- Senile brain amyloidosis
- Senile degeneration of brain
- Wilson's disease

**eFigure 2: Example of SNOMED expression generated for “forgetful”.**

Patient's husband states that patient has been more forgetful lately.

| | **SNOMED CT** |
|---|---|
| When? | 408731000\|Temporal context\|=6493001\|***Recent*** |
| What? | 246090004\|Associated finding\|=55533009\|***Forgetful*** |
| Who? | 408732007\|Subject relationship context\|=410604004\|***Subject of record*** |
| Present? | 408729009\|Finding context\|=410515003\|***Known present*** |
